# Reduction and Functional Exhaustion of T Cells in Patients with Coronavirus Disease 2019 (COVID-19)

**DOI:** 10.1101/2020.02.18.20024364

**Authors:** Bo Diao, Chenhui Wang, Yingjun Tan, Xiewan Chen, Ying Liu, Lifen Ning, Li Chen, Min Li, Yueping Liu, Gang Wang, Zilin Yuan, Zeqing Feng, Yuzhang Wu, Yongwen Chen

## Abstract

**BACKGROUND:** The outbreak of coronavirus disease 2019 (COVID-19) caused by severe acute respiratory syndrome coronavirus 2 (SARS-CoV-2) has posed great threat to human health, which has been declared a public health emergency of international concern (PHEIC) by the WHO. T cells play a critical role in antiviral immunity but their numbers and functional state in COVID-19 patients remain largely unclear.

**METHODS:** We retrospectively reviewed the counts of total T cells, CD4^+^, CD8^+^ T cell subsets, and serum cytokine concentration from inpatient data of 522 patients with laboratory-confirmed COVID-19, admitted into two hospitals in Wuhan from December 2019 to January 2020, and 40 healthy controls, who came to the hospitals for routine physical examination. In addition, the expression of T cell exhaustion markers PD-1 and Tim-3 were measured by flow cytometry in the peripheral blood of 14 COVID-19 cases.

**RESULTS:** The number of total T cells, CD4^+^ and CD8^+^ T cells were dramatically reduced in COVID-19 patients, especially among elderly patients (⩾60 years of age) and in patients requiring Intensive Care Unit (ICU) care. Counts of total T cells, CD8^+^T cells or CD4^+^T cells lower than 800/μL, 300/μL, or 400/μL, respectively, are negatively correlated with patient survival. Statistical analysis demonstrated that T cell numbers are negatively correlated to serum IL-6, IL-10 and TNF-α concentration, with patients in decline period showing reduced IL-6, IL-10 and TNF-α concentrations and restored T cell counts. Finally, T cells from COVID-19 patients have significantly higher levels of the exhausted marker PD-1 as compared to health controls. Moreover, increasing PD-1 and Tim-3 expression on T cells could be seen as patients progressed from prodromal to overtly symptomatic stages, further indicative of T cell exhaustion.

**CONCLUSIONS:** T cell counts are reduced significantly in COVID-19 patients, and the surviving T cells appear functionally exhausted. Non-ICU patients, with total T cells, CD8^+^T cells CD4^+^T cells counts lower than 800/μL, 300/μL, and 400/μL, respectively, may still require aggressive intervention even in the immediate absence of more severe symptoms due to a high risk for further deterioration in condition.

## Introduction

In December 2019, a series of acute respiratory illness were reported in Wuhan, Hubei Province, China.^1,2^ A novel coronavirus, initially named severe acute respiratory syndrome coronavirus 2 (SARS-CoV-2), was identified as the cause of this disease by the Chinese Center for Disease Control and Prevention (CDC).^3^ This disease, now designated as coronavirus disease 2019 (COVID-19) by the WHO, rapidly spread to other cities of China, and has become a public health emergency of international concern (PHEIC) following its global spread. COVID-19 is clinically manifests as fever, cough, muscle pain, fatigue, diarrhea and pneumonia, and can cause death in severe cases.^4-6^ Up through February 18, 2020, China has reported 72436 cases of confirmed COVID-19 and 1868 fatalities.^7^

Since an effective immune response against viral infections depends on the activation of cytotoxic T cells that can clear infection by killing virus-infected cells,^8^ boosting the numbers and function of T cells in COVID-19 patients is critical for successful recovery. A recent study reported that the 82.1% of COVID-19 cases displayed low circulating lymphocyte counts.^4-6^ However, the factors which might cause the reduction in count, and the activation status of T cells in COVID-19 patients, remain uninvestigated. We retrospectively analyze here the clinical data from 522 cases of COVID-19 who were admitted into the General Hospital of Central Theatre Command and Hanyang Hospital in Wuhan from December 2019 to January 2020. We also compared the expression of exhaustion markers PD-1 and Tim-3 on the surface of CD4^+^ and CD8^+^ T cells from COVID-19 and healthy controls. Our results thus provide a preliminary demonstration of T cell exhaustion during COVID-19 infection and suggest that more aggressive early intervention may be required in patients with low T lymphocyte counts.

## Methods

### Patients

Medical records from 522 patients (aged from 5 days to 97 years) with confirmed COVID-19 and admitted into the General Hospital of Central Theatre Command or Hanyang Hospital in Wuhan from December 2019 to January 2020, and 40 healthy people (aged from 2 to 62 years), who came to the hospitals for routine physical examination, were collected and retrospectively analyzed. Diagnosis of COVID-19 was based on the New Coronavirus Pneumonia Prevention and Control Program (5th edition) published by the National Health Commission of China.^9^ All the patients were laboratory-confirmed positive for SARS-CoV-2 by use of quantitative RT-PCR (qRT-PCR) of throat swab samples. This study was approved by the National Health Commission of China and Ethics Commission of General Hospital of Central Theatre Command ([2020]-004-1) and Hanyang Hospital (20200217). Written informed consent was waived by the Ethics Commission of the designated hospital for emerging infectious diseases.

### Definitions

The classification of clinical types, which consist of mild/moderate/severe/critical, was based on the New Coronavirus Pneumonia Prevention and Control Program (5^th^ edition) published by the National Health Commission of China.^9^ Within the cohort analyzed 43 were admitted to the intensive care unit (ICU), because they required high-flow nasal cannula or higher-level oxygen support measures to correct hypoxaemia. Hypoxaemia was defined as arterial oxygen tension (PaO2) over inspiratory oxygen fraction (FIO2) of less than 300 mm Hg or arterial oxygen saturation of 93% or lower. According to the staging of infectious disease,^10^ the prodromal period is a phase in which the host begins to experience general signs and symptoms. The illness period (overtly symptomatic period) is a phase in which the signs or symptoms of disease are most obvious and severe, with positive laboratory findings and chest images. For ICU patients, ICU period is a phase in which the **symptoms are most obvious and severe** The decline period is a phase in which the clinical symptoms begin to decline, laboratory findings and chest images improve, and arterial oxygen saturation to return to the normal.

### Data collection

We reviewed clinical records, nursing records, laboratory findings, and chest x-rays or CT scans for all the patients and physical examination records of the 40 healthy people. All information was obtained and curated with a customised data collection form. Three investigators (C Wang, Z Fen and Y Chen) independently reviewed the data collection forms to verify data accuracy.

### Sample collection and flow cytometric analysis

Peripheral blood samples from 14 patients and 3 healthy volunteers were simultaneously processed in the Central Lab of General Hospital of Central Theatre Command to isolate peripheral blood mononuclear cells (PBMCs) for further testing. The peripheral blood was supplemented with anticoagulants and PBMCs were harvested by density gradient centrifugation. Isolated PBMCs were stained with a BD multitest IMK Kit (Cat340503, BD Biosciences) for analyzing the frequency and cell number of total T, CD4+ T,CD8+ T, B and NK in healthy controls and patients. The exhaustion of T cells was detected using human CD4-percp (RPA-T4, Biolegend), CD8-APC (SK1, BD Biosciences), CD8-PE (SK1, Biolegend), PD-1-PE (EH12.2H7, Biolegend) and TIM-3-FITC (F38-2E2, Biolegend) antibodies. After being stained, the cells were measured by flow cytometry on an LSR Fortessa Cell Analyzer (BD Biosciences) and data analyzed using the FolwJo software (TreeStar). All experimental procedures were completed under biosafety level II plus condition.

### Statistical analysis

Statistical analyses were performed using GraphPad Prism version 8.0 (GraphPad Software, Inc., San Diego, CA, USA). Continuous variables were directly expressed as a range. Categorical variables were expressed as numbers/NUMBERS (%). p values are from χ2, non-paired t test or paired t test.

## Results

### 1. Decreasing the numbers of total T cells, CD4^+^ and CD8^+^ subsets in COVID-19 patients

From our retrospective analysis of 522 patients, 499 cases had lymphocyte count recorded. 75.75% (359/499), 75.95% (379/499) and 71.54% (357/499) patients had remarkably low total T cell counts, CD4^+^ and CD8^+^ T cell counts, respectively. Among milder patients in the Non-ICU group, the median value of total T cells, CD4^+^ and CD8^+^ T cell counts are 652, 342 and 208, respectively, the median value decreased to 261, 198 and 64.3, respectively, in the ICU group. (**Figure 1A**). The counts of total T cells, CD4^+^ and CD8^+^ T cells were significantly lower in ICU patients than Non-ICU cases (**Figure 1B**). These patients were further categorized into three groups based on age (<20 years old, 20∼60 years and ≥60 years), and an age-dependent reduction of T cell numbers was observed in COVID-19 patients, with the lowest T cells numbers found in patients ≥60 years old (**Figure 1C**), suggesting a potential cause for increased susceptibility in elderly patients.

**Figure 1.**
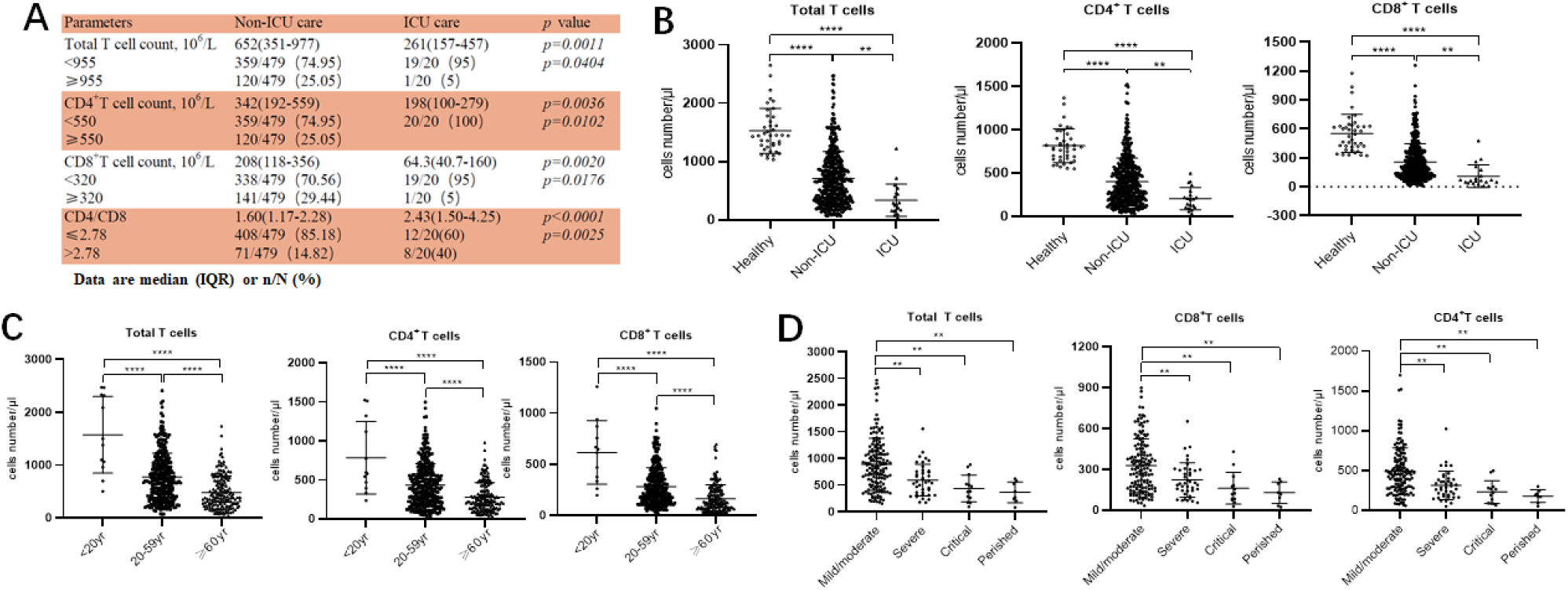
Reducing T cell numbers in COVID-19 patients. **A**. Demographics of T cells in patients; **B**. T cell numbers in different groups; **C**. T cell numbers in patients of different ages; **D**. T cell count in Non-ICU care patients with different clinical outcomes.\*\**p*<0.01, \*\*\**p*<0.001 and \*\*\*\**p*<0.0001.

We next retrospectively reviewed T cell numbers in 212 cases from Non-ICU patients within one center (the General Hospital of Central Theatre Command). The Non-ICU patients were further divided into four groups based on clinical outcomes. Among these patients, 151 cases are mild/moderate, 40 cases are severe, while 13 cases in critical condition, and 8 perished occurred. Statistical analysis showed that T cell numbers including total T cells, CD4^+^ and CD8^+^ T cells in severe, critical and perished groups are significantly lower than in the mild/moderate group. Most importantly, the numbers of total T cell, CD8^+^T cells and CD4^+^ T cells in severe and perished groups are lower than 800/μL, 300/μL, or 400/μL, respectively (**Figure 1D**). This result suggests that aggressive interventions may be required for non-ICU patients even in the absence of more severe symptoms should their T cell counts fall below the critical threshold.

### 2. Negatively correlated between T cell numbers and cytokines in COVID-19 patients

The expression of angiotensin converting enzyme 2(ACE2), the predicted receptor of SARS-CoV-2 viruses, is absent on T cells,^11^ suggesting that the depressed T counts in COVID-19 patients mentioned above (**Figure 1**) was likely not caused by direct infection of T cells. We therefore examined the concentrations of serum cytokines, including TNF-α, IFN-γ, IL-2, IL-4, IL-6 and IL-10, from these COVID-19 patients to explore the influence of cytokine signaling. We only found the levels of TNF-α, IL-6 and IL-10 were significantly increased in infected patients, and statistical analysis illustrated that their levels in ICU patients are significantly higher than in Non-ICU patients (**Figure 2A**).

**Figure 2.**
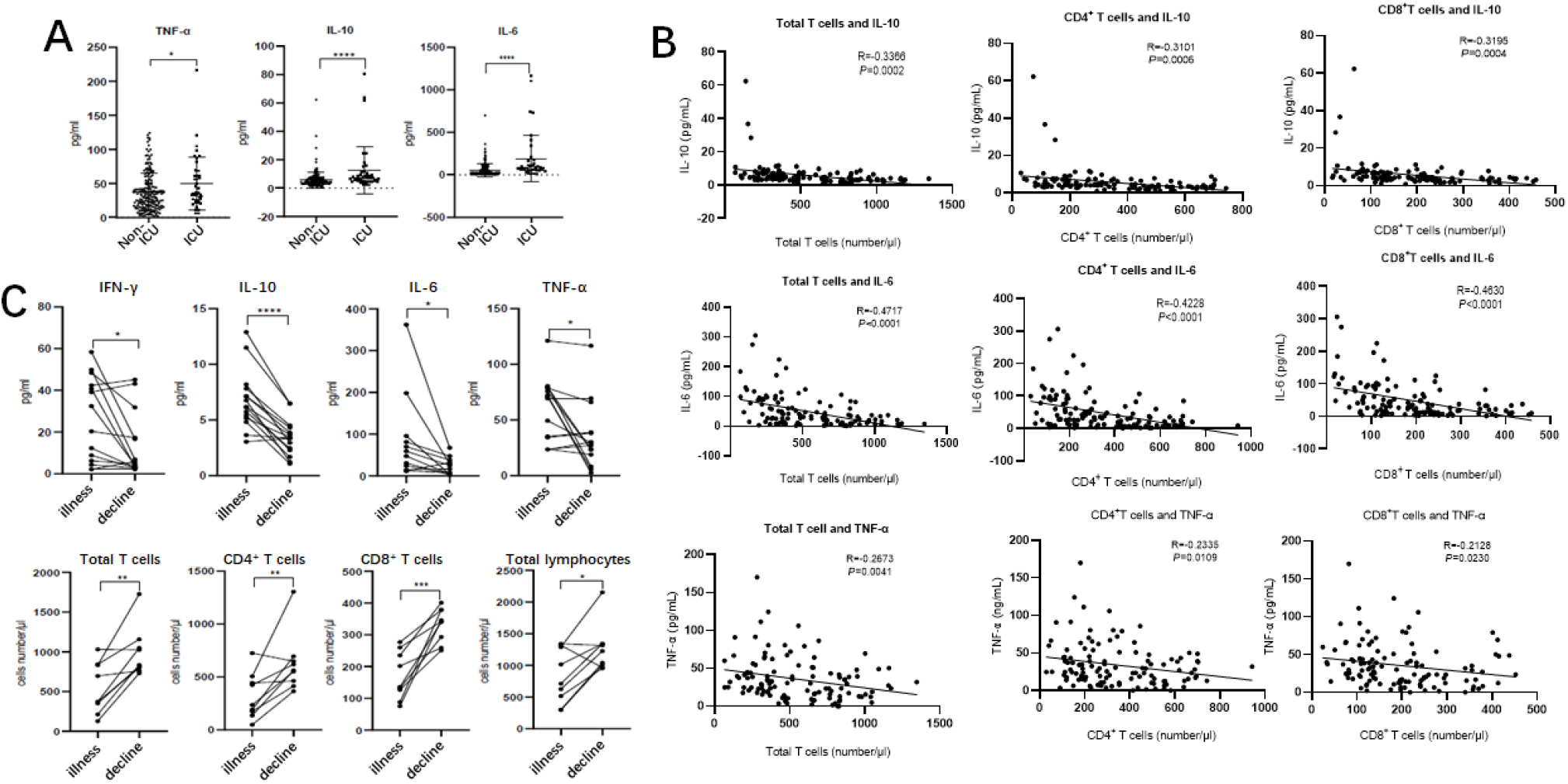
Cytokines and relative T cell numbers in COVID-19 patients. **A**. Demographics of cytokines in patients; **B**. Cytokine levels in different groups; **C**. The relativity of T cell numbers with cytokine levels; **D**. Dynamic profiles of cytokine levels and T cell numbers in Non-ICU care patients. \**p*<0.05, \*\**p*<0.01, \*\*\**p*<0.001 and \*\*\*\**p*<0.0001.

We next investigated the relationships between IL-10, IL-6, TNF-α and T cell count within Non-ICU patients. Interestingly, the concentration of these three cytokines was negatively correlated with total T cell counts, CD4^+^ counts, and CD8^+^ counts, respectively (**Figure 2B**). We subsequently summarized the follow up data of cytokine concentrations and T cell numbers in ten patients that were follower over the course of inpatient care. Interestingly, serum levels of IFN-γ, IL-10, IL-6 and TNF-α were significantly decreased in these patients in the decline period as compared with illness period, while counts of total T cells, CD4^+^, and CD8^+^ T cell subsets recovered during the decline period (**Figure 2C**). The phenomena suggests that the decrease of T cells seen in COVID-19 patients is likely the result of high serum concentration of TNF-α, IL-6 and IL-10 negatively regulating T cell survival or proliferation.

### 3. Enhancing T cell exhaustion in COVID-19 patients

Beyond changing in numbers during the course of infection, T cells may display limited function during prolonged infection as a result of exhaustion, which has been associated with the expression of some immune-inhibitory factors including PD-1, Tim-3 on cell surface.^12^ We therefore examined whether T cells in COVID-19 patients have exhaustion phenotypes. FACs analysis illustrated that, T cells from COVID-19 patients have markedly higher levels of PD-1 compared to healthy controls (Figure 3A). Furthermore, statistical analysis showed that the percentage of PD-1^+^CD8^+^ T cells from ICU patients was significantly higher than from both Non-ICU cases and healthy controls (**Figure 3B**), indicating that SARS-CoV-2 viruses induce T cell exhaustion in COVID-19 patients, particularly in those requiring ICU care.

**Figure 3.**
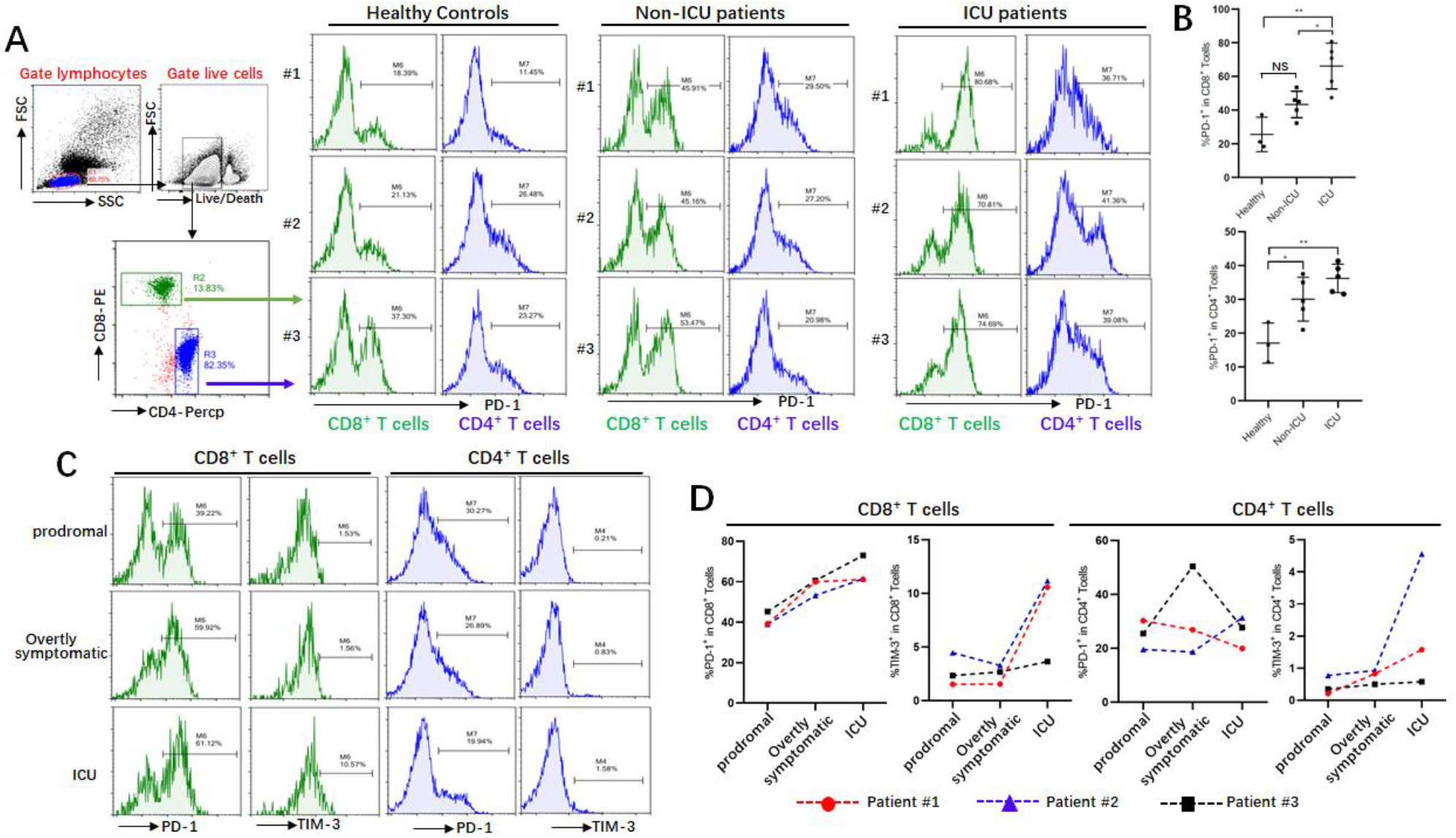
Exhaustion of T cells in COVID-19 patients. **A,B**. PD-1 expressions on T cells in different groups; **C, D**. Dynamic profile of PD-1 and TIM-3 expressions on T cells in 3 patients. NS: not significant, \**p*<0.05, \*\**p*<0.01.

Three patients were follow-up during inpatient care, and the expression of the exhausted markers including PD-1 and Tim-3 on surface of T cells during disease progress was detected. FACs showed that these patients have very low level of PD-1 and Tim-3 on CD8^+^ and CD4^+^ T cells in the prodromal stage, however, their levels on CD8^+^ T cells was increased in overtly symptomatic stages, and highest levels were seen in ICU period (**Figure 3C, D**). Similarly, higher levels of Tim-3 was observed on CD4^+^ T cells from patients who are in ICU stage, although enhancing the expression of PD-1 on CD4^+^ T cells was not obviously during disease progress (**Figure 3C, D**). These results demonstrated that T cells are exhaustion in COVID-19 patients during SARS-CoC-2 infection.

## Discussion

T cells play a vital role in viral clearance, with CD8^+^ cytotoxic T cells (CTLs) capable of secreting an array of molecules such as perforin, granzymes, and IFN-γ to eradicate viruses from the host.^13^ At the same time, CD4^+^ helper T cells (Th) can assist cytotoxic T cells and B cells and enhance their ability to clear pathogen.^14^ However, persistent stimulation by the virus may induce T cell exhaustion, leading to loss of cytokine production capability and reduced functions. ^15,16^ Earlier studies have been unclear regarding the numbers and function of T cells in COVID-19 patients, albeit with suggestions of depressed lymphocyte counts.^4,6^ In this report, we retrospectively reviewed the numbers of total T cells, CD4^+^, CD8^+^ T cell subsets in a total of 499 COVID-19 patients. In Non-ICU patients, we found that over 70.56% cases underwent decreased in the total T cells, CD4^+^ T cells and CD8^+^ T cells. However, in the ICU group, a total of 95% (19/20) patients showed a decrease in both total T cells and CD4^+^ T cells, and most importantly, all of the patients displayed decreases in CD8^+^ T cells. We also analyzed Non-ICU patients in greater detail, and found that aggressive intervention may be necessary to preempt the development of severe symptoms in patients with low T cell counts.

Cytokine storm is a phenomenon of excessive inflammatory reaction in which cytokines are rapidly produced in large amount in response to microbial infection. This phenomenon has been considered an important contributor to acute respiratory distress syndrome (ARDS) and multiple organ dysfunction syndrome (MODS).^17,18^ It has been also implicated in the setting of respiratory viral infections, such as SARS in 2002, avian H5N1 influenza virus infection in 2005 and H7N9 infection in 2013.^19-22^ Huang C et al. showed that the levels of IL-2, IL-7, IL-10, TNF-α, G-CSF, IP-10, MCP-1 and MIP-1A were significantly higher in COVID-19 patients.^4^ Consistent with this report, we here also found that the secretion of cytokines including TNF-α, IL-6 and IL-10 was increased in COVID-19 patients. Interestingly, the numbers of total T cells, CD4^+^ T and CD8^+^ T cells are negatively correlated to levels of TNF-α, IL-6 and IL-10, respectively (**Figure 2B**), suggesting these cytokines promote T cells decrease in COVID-19 patients.

TNF-α is a pro-inflammatory cytokine which can promote T cell apoptosis *via* interacting with its receptor, TNFR1, which expression is increased in aged T cells.^23,24^ Our current analysis demonstrated that patient over 60 years old have lower T cell numbers, indicating that TNF-α might be directly involved in inducing T cell loss in these patients. IL-6, when promptly and transiently produced in response to infections and tissue injuries, contributes to host defense through the stimulation of acute phase responses or immune reactions. Dysregulated and continual synthesis of IL-6 has been shown to play a pathological role in chronic inflammation and infection.^25,26^ Tocilizumab, a humanized anti-IL-6 receptor antibody, has been developed and approved for the treatment of rheumatoid arthritis (RA) and juvenile idiopathic arthritis.^27,28^ Moreover, tocilizumab has been shown to be effective against cytokine release syndrome resulting from CAR-T cell infusion against B cell acute lymphoblastic leukemia.^29^ Whether tocilizumab can restore T cell counts in COVID-19 patients by suppressing IL-6 signaling remains uninvestigated.

One interesting question is the source of these cytokine during COVID-19 infection. While previous studies have validated that the secretion of cytokines including IL-6, IL-10 and TNF-α are mainly from T cells, macrophages and monocytes etc, based on our results, we suggest that the secretion of these cytokines does not originate from T cells. However, the cytokine storm in turn may promote apoptosis or necrosis of T cells, and consequently leads to their reduction. Our previous work demonstrating that monocytes and macrophages can produce pro-inflammatory cytokine during murine hepatitis virus strain-3 infection,^30,31^ and whether SARS-CoV-2 also triggers cytokine release from monocytes and macrophages in COVID-19 patients need further investigation and such work is in progress in our hospital.

T cell exhaustion is a state of T cell dysfunction that arises during many chronic infections and cancer. It is defined by poor effector function, sustained expression of inhibitory receptors, and a transcriptional state distinct from that of functional effector or memory T cells.^32^ By FACs analysis, we found that both CD8^+^ T cells and CD4^+^ T cells have higher levels of PD-1 in virus infected patients, particularly when derived from ICU patients. Since these changes could also be observed in our longitudinal follow-up of several patients from prodromal to ICU care (**Figure 3**). IL-10, an inhibitory cytokine, not only prevents T cell proliferation, but also can induce T cell exhaustion. Important, blocking IL-10 function has been shown to successfully prevent T cell exhaustion in animal models of chronic infection.^33,34^ We demonstrate here that COVID-19 patients have very high levels of serum IL-10 following SARS-CoV-2 infection, while also displaying high levels of the PD-1 and Tim-3 exhaustion markers on their T cells, suggesting that IL-10 might be mechanistically responsible. The application of potent antiviral treatments to prevent the progression to T cell exhaustion in susceptible patients may thus be critical to their recovery. We have read with great interest the successful application of Remdesivir to curing a COVID-19 patient in the US, and to clinical trials indicates that it may have significant potential as such an antiviral.^35,36^

Taken together, we conclude that T cells are decreased and exhausted in patients with COVID-19. Cytokines such as IL-10, IL-6 and TNF-α might directly mediate T cell reduction. Thus, new therapeutic measures are needed for treatment of ICU patients, and may even be necessary early on to preempt disease progression in higher-risk patients with low T cell counts.

## Data Availability

For protection of patient privacy, all data used during the study only be provided with anonymized data.

